# Gene-based polygenic risk scores analysis of alcohol use disorder in African Americans

**DOI:** 10.1101/2022.01.19.22269532

**Authors:** Dongbing Lai, Tae-Hwi Linus Schwantes-An, Marco Abreu, Grace Chan, Victor Hesselbrock, Chella Kamarajan, Yunlong Liu, Jacquelyn L. Meyers, John I Nurnberger, Martin H. Plawecki, Leah Wetherill, Marc Schuckit, Pengyue Zhang, Howard J. Edenberg, Bernice Porjesz, Arpana Agrawal, Tatiana Foroud

**Author notes:** Corresponding Author Dongbing Lai, Ph.D.

## Abstract

Genome-wide association studies (GWAS) in admixed populations such as African American (AA) have limited sample sizes, resulting poor performance of polygenic risk scores (PRS). Based on the observations that many disease-causing genes are shared between AA and European ancestry (EA) populations, and some disease-causing variants are located within boundaries of these genes, we proposed a novel gene-based PRS method (PRS_gene_) by using variants located in these shared disease-causing genes. Using AA GWAS of alcohol use disorder (AUD) from the Million Veteran Program and EA GWAS of problematic alcohol use as the discovery datasets, we identified 858 variants from 410 genes that were AUD-related in both AA and EA. PRS_gene_ calculated using these variants were significantly associated with AUD in three AA cohorts (P-values: 7.61E-05-6.27E-03; Betas: 0.15-0.21) and outperformed PRS calculated using all variants (P-values: 7.28E-03-0.16; Betas: 0.06-0.18). PRS_gene_ was also associated with AUD in an EA cohort (P-value=0.02, Beta=0.11). In AA, individuals in the highest PRS_gene_ decile had an Odds Ratio of 1.76 (95% CI: 1.32-2.34) to develop AUD compared to those in the lowest decile. The 410 genes were enriched in 54 Gene Ontology biological processes, including ethanol oxidation and processes involving synaptic system, which are known to be AUD-related. Additionally, 26 genes were targets of drugs to treat AUD or other diseases, but may be repurposed to treat AUD. Our study demonstrated that our gene-based PRS had improved performance in evaluating AUD risk in AA and provided new insight into identification of AUD genes.

## INTRODUCTION

Alcohol use disorder (AUD) is one of the most common public health problems (1) and caused by both genetic and environmental factors. Estimates of the heritability of AUD ranges from 40% to 60% (2-4). Recently, multiple large-scale genome-wide association studies (GWAS) of AUD-related phenotypes have been published (5-7) and many variants associated with AUD have been identified. These GWAS reinforce the highly polygenic architecture underlying AUD, with many variants exerting small effects. Consequently, polygenic risk scores (PRS) have proven to be a strong approach for assessing AUD genetic liability beyond genome-wide significant variants (5, 7). For instance, in a recent study (Lai et al, accepted), individuals comprising the highest PRS decile were almost twice as likely to meet criteria for AUD relative to all others in a European ancestry sample, an estimate comparable to those published for first degree family history of AUD in national surveys (8, 9). However, PRS analysis of AUD in admixed populations, such as African American (AA), suffer from poor performance due to the smaller sample sizes of the discovery GWAS (5).

While increasing the discovery sample sizes is the ultimate way to improve PRS performance, sample sizes comparable to European ancestry populations (hundreds thousands to >1 million) for admixed populations will require extensive and strategic data collection. Studies have shown that many disease-causing genes are shared among different populations (10-13). Therefore, large-scale EA GWAS summary statistics can be leveraged to improve the performance of PRS in non-EA populations by increasing the overall discovery GWAS sample size. However, disease-causing variants may have different allele frequencies and effect sizes in different populations, and linkage disequilibrium (LD) patterns in admixed populations are more complicated than those in EA populations (11, 14-17). Methods utilizing the large EA GWAS and smaller non-EA GWAS to calculate PRS in non-EA populations aimed to address these challenges have been proposed, but their performance remains far from ideal (18-22).

Optimizing the selection of variants to include in PRS improves performance. Majority variants in the genome are likely not related to a particular condition and including them in PRS calculations will reduce performance by introducing noises. Ideally, only variants that act on disease-causing genes should be used in calculating PRS. However, most of these genes and variants remain to be discovered. Since many disease-causing genes are shared across different populations (10-13), GWAS from different populations provide another way to identify these genes and variants. For instance, if a gene is nominally associated (e.g. P-values <0.05) with a disease in both EA and non-EA populations, then it is likely to be a shared disease-causing gene across populations. Furthermore, for these disease-causing genes, it is highly likely that there is at least one disease-causing variant located within the gene boundaries, e.g. non-synonymous mutations that change gene products or 3’ UTR variants that alter gene expression levels (23, 24). Additionally, regulatory variants that are close to a gene may be tagged by variants located within the gene boundaries due to LD. Therefore, variants located in gene boundaries that are nominally associated with a disease in both EA and non-EA populations are likely to be disease-causing variants or in LD with disease-causing variants. Based on these observations, we proposed a novel method aimed at further enhancing the performance of PRS in admixed population, referring to this method as gene-based PRS. We first used EA GWAS and non-EA GWAS to identify shared disease-causing genes, then used variants located within these genes’ boundaries to calculate PRS. While variants acting distally are not included in this calculation, by excluding majority variants in the genome that are unlikely to be related to a disease, the signal to noise ratio theoretically increases and thus the performance of PRS is expected to improve. Further, if the gene-based PRS are significantly associated with a disease, it becomes more likely that some or most of these included genes are truly disease-causing, despite not previously being implicated by GWAS or gene-based genome-wide association studies like those using MAGMA (25). Thus gene-based PRS provide another opportunity to pinpoint the disease-causing genes with sub-threshold P-values.

In this study, we examined the performance of the proposed gene-based PRS in evaluating the risk of AUD in AA and EA populations. Using summary statistics from an EA GWAS of problematic alcohol use (EA-PAU) (7) and an AA GWAS of AUD from the Million Veteran Program (AA-AUD) (5), the largest GWAS of AUD-related phenotypes to date in EA and AA populations respectively as the discovery GWAS, we calculated gene-based PRS (PRS_gene_) to evaluate AUD risk in three AA cohorts: the Collaborative Study on the Genetics of Alcoholism (COGA) (26), Study of Addiction: Genetics and Environment (SAGE)(27), Alcohol Dependence GWAS in European and African Americans (YalePenn, AA participants only) (28), as well as one EA cohort from the Indiana Biobank (https://indianabiobank.org/). We compared the performance of PRS_gene_ with PRS calculated using all variants (PRS_all_). Then, for genes included in PRS_gene_ calculations, we performed gene ontology enrichment analysis to test whether they were enriched in biological processes known to be related to AUD or other pathways that could provide novel insight into AUD etiology. We also searched a publically available drug target database (29) to evaluate whether these genes were potential drug targets for AUD treatment, or drug targets for treatment of other diseases but may be repurposed to treat AUD.

## METHODS

### Discovery datasets

The EA-PAU was a meta-analysis of problematic alcohol use (N=435 563) (7) that comprised an AUD GWAS of the Million Veteran Program Phase I (5) and Phase II data, an alcohol dependence GWAS from the Psychiatric Genomics Consortium (30), and a GWAS of scores from the problem subscale of the Alcohol Use Disorder Identification Test (AUDIT items 4 to 10) in the UK Biobank (6). AA-AUD (N=56 648) (5) served as the discovery sample for AA target samples. Across both discovery GWAS, A/T or C/G variants were excluded to avoid strand ambiguity. As we were focusing on AUD-causing variants shared between EA and AA, only variants having the same direction of effects in both the EA-PAU and AA-AUD were included. Both GWAS summary statistics were downloaded from the database of genotypes and phenotypes (dbGaP: phs001672.v3.p1, https://www.ncbi.nlm.nih.gov/projects/gap/cgi-bin/study.cgi?study_id=phs001672.v3.p1).

### Target datasets

AA individuals were drawn from 3 sources: COGA (N=3 375), SAGE (N=930), and YalePenn (N=2 010). COGA is a family cohort, in which alcohol dependent probands and their family members from inpatient and outpatient AD treatment facilities in seven sites were invited to participate. COGA also recruited community comparison families from a variety of sources in the same areas (26, 31). The study was approved by Institutional review boards from all sites. Every participant provided informed consent. The Semi-Structured Assessment for the Genetics of Alcoholism (SSAGA) was administered to individuals 18 or over and the child version of the SSAGA was used for those younger than 18 (32, 33). SAGE (phs000092.v1.p1, https://www.ncbi.nlm.nih.gov/projects/gap/cgi-bin/study.cgi?study_id=phs000092.v1.p1) and YalePenn (phs000425.v1.p1, https://www.ncbi.nlm.nih.gov/projects/gap/cgi-bin/study.cgi?study_id=phs000425.v1.p1) were downloaded from dbGaP. Since COGA had more phenotypic information, if a sample in the COGA dataset was also in SAGE and/or YalePenn, they were only analyzed as part of the COGA data. SAGE and YalePenn are mix of related and unrelated individuals, although most are unrelated. Only AA samples from COGA, SAGE, and YalePenn were used. Across all three datasets, AUD was defined as meeting lifetime criteria for DSM-IV alcohol dependence (34) or DSM-5 alcohol use disorder (35). All other individuals were considered as controls.

EA individuals were drawn from one source (i.e. Indiana Biobank). The gene-based PRS was calculated using variants located in AUD genes implicated in both AA and EA. Consequently, it should be applicable to both populations. To test this proposition, we included a European ancestry dataset from the Indiana Biobank (IB: https://indianabiobank.org/). IB is a state-wide collaboration that provides centralized processing and storage of specimens that are linked to participants’ electronic medical information via Regenstrief Institute at Indiana University. All IB individuals included in this study are unrelated. AUD in IB was diagnosed based on ICD9 (303 and 305.0) and ICD10 (F10) codes. Individuals not diagnosed as AUD and without AUD associated conditions such as alcohol-associated pancreatitis were defined as controls. As some EA samples of COGA and SAGE data were part of EA-PAU, they were not included as target EA samples.

### Genotype data processing and Imputation

Detailed information about COGA, SAGE, and YalePenn data processing has been reported (36-38). Briefly, a common set of high quality (minor allele frequency (MAF) >10%, missing rate <2%, Hardy-Weinberg Equilibrium (HWE) P-values >0.001) and independent (defined as R^2^ <0.5) variants (N=24 135) was used to identify duplicate samples among different cohorts and confirm the reported family structures using PLINK (39, 40). Family structures were updated if needed. The same set of common variants was also used to estimate the principal components (PCs) of population stratification using Eigenstrat (41) with 1000 Genomes data (Phase 3, version 5, NCBI GRCh37) as the reference panel. Each cohort was imputed to 1000 Genomes by using SHAPEIT2 (42) followed by Minimac3 (43). Before imputation, variants with A/T or C/G alleles, missing rates >5%, MAF <3%, and HWE P-values <0.0001 were excluded. Imputed variants with R^2^ >=0.30 and MAF >=1% were included in all analyses. As previously reported, IB samples were genotyped using Illumina Infinium Global Screening Array (GSA, Illumina, San Diego, CA) by Regeneron (Tarrytown, NY)(44). Variants with missing rate >5%, MAF <1%, HWE P-value <1E-10 among cases and 1E-6 in controls were excluded. Population stratification was then estimated using the SNPRelate package (45) from Bioconductor (46). IB data were also imputed to 1000 Genomes using the Michigan Imputation Server (https://imputationserver.sph.umich.edu/index.html#!pages/home) (43). Imputed variants with R^2^ <0.30 and MAF <1% were excluded.

### PRS calculation

We used PRS-CSx, a recently developed method designed for cross-ethnic polygenic prediction and showed better performance when compared with other methods in simulation studies and real data analysis (22). The posterior effect size of each variant was estimated via a Bayesian regression framework using continuous shrinkage priors. African and European samples from the 1000 Genomes project were used as the LD reference panels. PRS-CSx can estimate posterior effect sizes of AA only, EA only, and meta-analysis of EA-PAU and AA-AUD. Since we focused on AUD-causing variants implicated in both AA and EA, meta-analysis posterior effect sizes were used. Only variants located within gene boundaries (defined as ±1 kb from the transcription start and end sites), having P-values <0.05 in both EA-PAU and AA-AUD (i.e., at least showing marginal associations), and having the same directions of effects were included in PRS calculations (referred to as PRS_gene_). We also used all variants to calculate PRS (PRS_all_) for comparison purposes. PLINK (39, 40) was used to calculate PRS using the posterior effect sizes estimated by the PRS-CSx. PRS_gene_ and PRS_all_ were standardized as mean=0 and standard deviation=1 in AA (all three datasets combined) and EA target dataset separately.

### Statistical analysis

As COGA, SAGE, and YalePenn included related individuals, generalized linear mixed models were used with a random effect to adjust for family relationships. For IB, which is a cohort of unrelated individuals, logistic regression models were used. We also stratified individuals based on PRS deciles and compared each to the bottom decile. Since the sample sizes in COGA, SAGE, and YalePenn are small to moderate (therefore having insufficient sample sizes in each decile), we combined all three cohorts for the stratified analyses. For all models, sex and the first 10 PCs were included as covariates. For the combined analysis of COGA, SAGE, and YalePenn data, a cohort indicator (i.e. COGA, SAGE, or YalePen) was also included. Associations with P-values <0.05 across all 3 cohorts were considered statistically significant for PRS_gene_ and PRS_all_.

### Gene Ontology enrichment analyses, searching GWAS catalog and potential drug target genes

For genes included in calculating PRS_gene_, we performed gene ontology enrichment analysis using PANTHER (released 2021-01-24) (47) implemented in the Gene Ontology (GO) Resource (http://geneontology.org/, released 2021-08-18). We focused on GO Biological Processes (GOBPs), which were compiled by multiple molecular activities. We also searched the GWAS catalog (https://www.ebi.ac.uk/gwas/) (48) to check whether these genes had been previously implicated in GWAS of AUD-related phenotypes. Lastly, we checked whether these genes could be potential drug targets by searching the gene list for targets of FDA-approved drugs as well as those in current clinical trial investigations, compiled by Wang et al (29) derived from the Informa Pharmaprojects database (https://pharmaintelligence.informa.com/products-and-services/data-and-analysis/pharmaprojects).

## RESULTS

Samples used in this study are summarized in **Table 1**. In all cohorts, about 60% of cases were males while <42% of controls were males.

**Table 1:** Sample summary.

| Population | cohort | # case (%male) | # control (%male) | # total | # families |
| --- | --- | --- | --- | --- | --- |
| AA | All* | 2,786 (60.80) | 3,529 (39.27) | 6,315 | 3,322 |
|  | COGA | 875 (62.06) | 2,500 (41.60) | 3,375 | 590 |
|  | SAGE | 387 (59.17) | 543 (37.02) | 930 | 869 |
|  | YalePenn | 1,524 (60.50) | 486 (29.84) | 2,010 | 1,863 |
| EA | IB | 539 (62.15) | 3,515 (40.40) | 4,054 | 4,054 |

\*: COGA, SAGE, and YalePenn combined.

PRS-CSx estimated posterior meta-analysis effect sizes for 1 126 428 variants and they were used to calculate PRS_all_. Among these variants, 858 variants with P-values <0.05 in both the EA-PAU and AA-AUD (**Table S1**) that were located within gene boundaries (410 genes, **Table S2**) were used to calculate PRS_gene_. As shown in **Table 2**, for AA cohorts, both PRS_gene_ and PRS_all_ had P-values<0.05 in all cohorts except PRS_all_ for COGA. Effect sizes ranged from 0.15-0.21 for PRS_gene_ and 0.06-0.18 for PRS_all_. In EA, 847 of 858 variants (**Table S1**), and 1 061 130 of 1 126 428 variants were present in IB after QC; both PRS_gene_ and PRS_all_ had P-values<0.05 (PRS_gene_ Beta=0.11, SE=0.02; PRS_all_ Beta=0.34, SE=0.05). Overall, PRS_gene_ outperformed PRS_all_ in AA but the opposite was observed in EA.

**Table 2:** Associations between AUD and PRS_gene,_ PRS_all_ in AA and EA.

| Population | cohort | PRS<sub>gene</sub> |  |  |  | PRS<sub>all</sub> |  |  |  |
| --- | --- | --- | --- | --- | --- | --- | --- | --- | --- |
|  |  | Beta | SE | P-value | # Variants | Beta | SE | P-value | # Variants |
| AA | All<sup>*</sup> | 0.17 | 0.03 | **3.55E-08** | 858 | 0.12 | 0.03 | **9.42E-05** | 1,126,428 |
|  | COGA | 0.15 | 0.04 | **9.67E-04** | 858 | 0.06 | 0.04 | 0.16 | 1,126,428 |
|  | SAGE | 0.18 | 0.07 | **6.27E-03** | 858 | 0.18 | 0.07 | **0.01** | 1,126,428 |
|  | YalePenn | 0.21 | 0.05 | **7.61E-05** | 858 | 0.17 | 0.06 | **7.28E-03** | 1,126,428 |
| EA | IB | 0.11 | 0.05 | **0.02** | 847 | 0.34 | 0.05 | **2.35E-21** | 1,061,130 |

\*: COGA, SAGE, and YalePenn combined. PRS<sub>gene</sub>: PRS calculated using variants located in genes associated with AUD in both AA and EA. PRS<sub>all</sub>: PRS calculated using all variants.
Significant P-values are in bold.

The association between PRS and AUD increased from the bottom decile (1^st^ decile) to the top decile (10^th^ decile) (**Figure 1)**. Using the bottom decile as the reference, all except the 2^nd^ and 3^rd^ deciles were associated with the increased risk for AUD (ORs: 1.37-1.76. **Table 3**) after adjusting for covariates.

**Table 3:** Odds Ratios when comparing each PRS_gene_ decile with the bottom decile in AA.

| Decile | OR | OR 95%CI | P-value |
| --- | --- | --- | --- |
| 10 | 1.76 | 1.32-2.34 | **1.03E-04** |
| 9 | 1.72 | 1.29-2.29 | **2.05E-04** |
| 8 | 1.52 | 1.15-2.00 | **2.93E-03** |
| 7 | 1.64 | 1.27-2.12 | **1.49E-04** |
| 6 | 1.43 | 1.09-1.87 | **0.01** |
| 5 | 1.37 | 1.05-1.79 | **0.02** |
| 4 | 1.48 | 1.13-1.93 | **4.55E-03** |
| 3 | 1.22 | 0.91-1.62 | 0.18 |
| 2 | 1.11 | 0.85-1.45 | 0.44 |

Note: Significant P-values are in bold.

**Figure 1:**
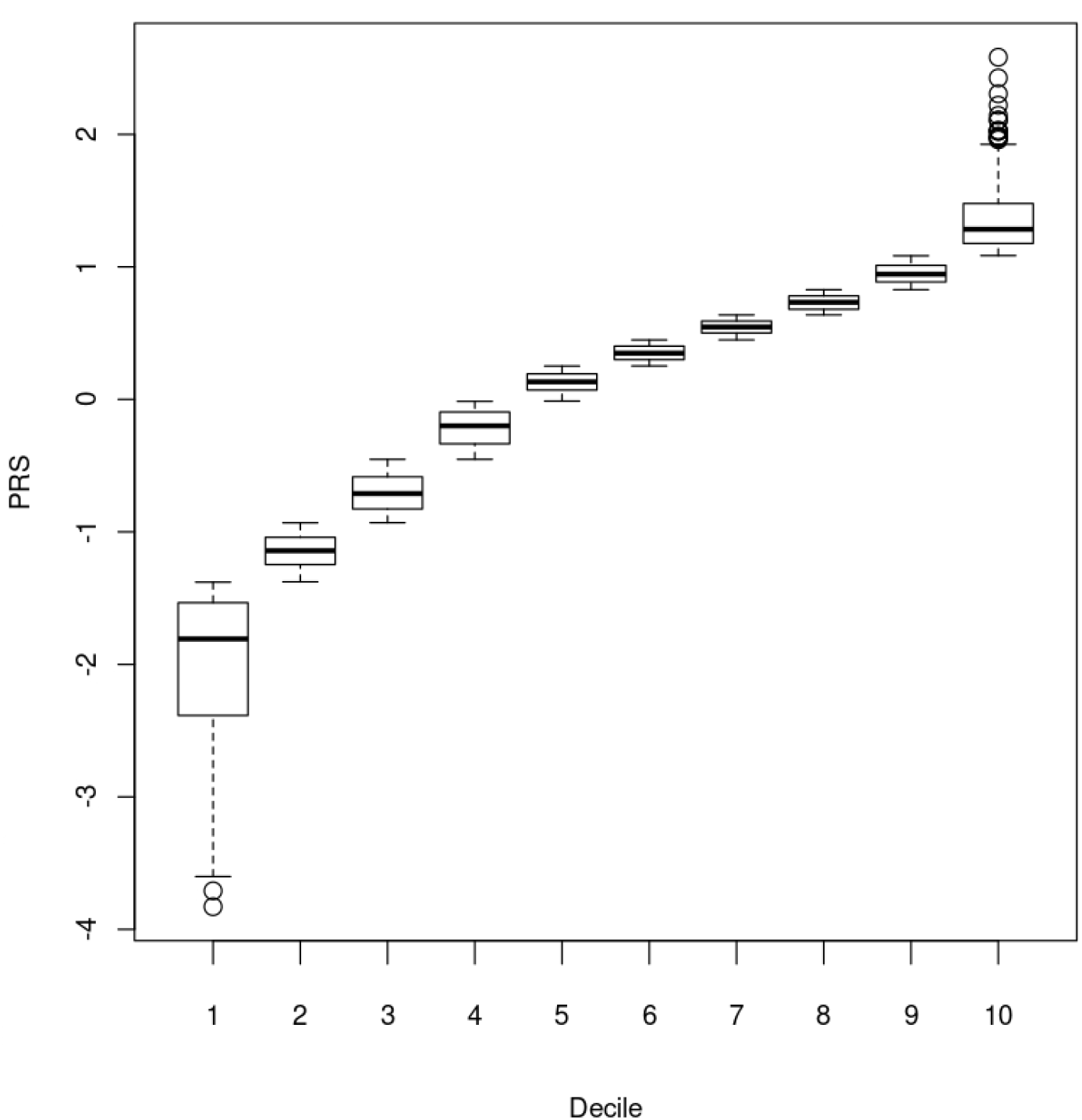
Distributions of PRS_gene_ in each decile in AA.

Of the 410 genes, 353 were uniquely mapped to the GO database. The unmapped genes were non-coding RNAs, anti-sense RNAs, pseudo-genes, and read-throughs. Fifty four GOBPs had false discovery rate (FDR) P-values <0.05, including ethanol oxidation, synaptic signaling, synapse organization, synaptic plasticity, startle response, neurogenesis, nervous system development, learning or memory, protein metabolic process, cell adhesion, cell development, cell junction organization, movement of cell or subcellular component, cell-cell signaling, regulation of signaling, etc. (**Table S3**).

Only 47 genes were identified in previous GWAS of AUD-related phenotypes (**Table S2**). Twenty six genes were targets of drugs approved by FDA or in clinical trials. Among them, four (*DRD2, PDE4B, GRM5*, and *SLC6A9*; the first two were reported by previous GWAS of AUD-related phenotypes) were drug target genes for AUD treatment (**Table S4**); for those 22 genes that were targets of drugs to treat diseases other than AUD, 21 were involved in the significant GOBPs identified and five (*EIF4E, ESR1, MAPT, METAP1, and TNKS*) were reported by previous GWAS of AUD related phenotypes (**Table S2**).

## DISCUSSION

In this study, we found that gene-based PRS (PRS_gene_) calculated using 858 variants of 410 genes were significantly associated with AUD in both AA and EA, and outperformed the PRS calculated using all variants (PRS_all_) in AA. Compared to the bottom decile, those at the top PRS_gene_ decile were nearly twice as likely to be AUD cases (OR=1.76) in AA. These 410 genes were enriched in 54 GOBPs, and many of them are likely to be AUD-related. Additionally, four genes were targets of drugs in Phase II or III clinical trials to treat AUD; 22 genes were targets of drugs approved by FDA or in clinical trials to treat other diseases but may be repurposed to treat AUD. Together, these findings showed that a biologically meaningful polygenic signal can be characterized in non-European ancestry individuals by leveraging methods that focus on intragenic signals with concordant directions of effects across ancestries. Further, the process identified drugs already under development that should be evaluated for their potential in AUD.

To improve the performance of PRS, more disease-causing variants should be included and unrelated variants should be excluded. AUD is caused by many genes with small effects and in GWAS of AUD, due to the large number of variants tested, many variants that are unrelated to AUD show some degree of association (e.g. P-values <0.05) purely by chance (i.e., false positives). If sample sizes are large (e.g. hundreds of thousands of participants or more), while a majority of AUD-related variants are still not genome-wide significant, they usually have smaller P-values than those false positives and can still contribute to the calculation of PRS. However, when the discovery GWAS sample sizes are small to moderate, the discrimination between AUD-related and unrelated variants narrow. This may lead to a reduction in PRS performance. Using large-scale EA discovery GWAS could mitigate this problem, but the improvement is limited even with sophisticated statistical methods (22). Using our gene-based PRS strategy, we focused on genes with nominal association in both EA and AA then used variants located in these genes to calculate PRS_gene_. By excluding most unrelated variants, the chance of selecting true AUD-related variants was greatly increased; and as shown in our study, resulted in superior performance in all our AA cohorts. Thus, we conclude that this strategy can be used to improve the performance of PRS when the discovery GWAS sample sizes are not sufficiently large, notable in admixture populations and other groups that have been underrepresented in GWAS studies to date.

While PRS_gene_ outperformed PRS_all_ in AA, the opposite was observed in EA. This is expected for the following reasons. First, many GWAS findings, such as variants in *KLB* and *GCKR*, which reached genome-wide significance in EA, had P-values >0.05 in AA (i.e. these genes may not be AUD-related in AA for some unknown mechanisms, or variants act on these genes in AA have not been identified), therefore, they were not included in calculating PRS_gene_ but were used in calculating PRS_all_ in EA. Second, even within genes that have shown associations with AUD in both AA and EA, different causal variants may have been important in each ancestral group. One example is rs2066702 in the *ADH1B* gene. While relatively common in AA individuals (MAF=0.18), the variant is rare in EA individuals (MAF=0.002) (https://www.ncbi.nlm.nih.gov/snp/rs2066702?vertical_tab=true#frequency_tab). This is the only variant selected in *ADH1B* in calculating PRS_gene_, resulting in no contribution of *ADH1B* when calculating PRS_gene_ in EA individuals from the Indiana Biobank. However, for PRS_all_, multiple common EA variants in *ADH1B* (e.g. rs2066701, rs1042026, and rs2075633) were included, thus increasing the performance of PRS_all_. Third, we limited inclusion to variants within gene boundaries and this may affect AA and EA disproportionately. For example, rs1229978, which is located between *ADH1B* and *ADH1C*, is much more common in EA (MAF=0.39) than in AA (MAF=0.15) (https://www.ncbi.nlm.nih.gov/snp/rs1229978?vertical_tab=true); therefore, not including this variant in PRS calculations had larger impact in EA than in AA. Nevertheless, the significance of PRS_gene_ in both AA and EA suggested that most of these genes are AUD-related in these two populations.

Most variants identified in GWAS are typically outside gene boundaries, therefore, post-GWAS functional studies (in silico or in wet lab) are usually performed to identify true AUD-associated genes. Our gene-based PRS provided another strategy to pinpoint these AUD associated genes. By focusing on variants within gene boundaries that were nominally associated with AUD in AA and EA, if PRS calculated using these variants were significantly associated with AUD, then genes that host these variants were likely to be AUD-associated. In this study, started with >1M variants, we ended up using 858 variants located in 410 genes to calculate PRS_gene_. The significant results of PRS_gene_ in both AA and EA indicated that most of these 410 genes contributed to the association signal and were likely to be AUD-related. There exist multiple gene-based methods such as MAGMA (25) to consolidate genome-wide variant-level signals, however, these methods are hypothesis free and can still suffer an attenuated but harsh multiple testing correction. As a result, most genes will not meet genome-wide significance (e.g. in our study, 363 of those 410 genes were not mentioned by any GWAS of AUD) and additional studies are still needed to differentiate the true disease-causing genes from falsely identified ones. Furthermore, gene-based methods like MAGMA test the same GWAS samples, while our gene-based PRS test independent samples, increasing replicability.

More than half of the 410 genes (244) were involved in 54 significant GOBPs. As expected, ethanol oxidation was among them and four genes (*ADH1B, ADH1C, ADH4*, and *ADH5*) were involved. Compromised executive functioning (i.e., neuroadaptation) is one of the major mechanisms contributing to AUD (49) and not surprisingly, several significant GOBPs related to synaptic systems (synaptic signaling, synapse organization, synaptic plasticity, startle response) were identified (46 genes). Although the role of the synaptic system in AUD is well-established (49), however, only nine genes (*CSMD1, DCC, DRD3, EIF4E, ERC2, LINGO2, MAPT, NRXN2*, and *TENM2*) were implicated in previous GWAS of AUD related phenotypes. We also found significant GOBPs related to learning and memory (27 genes), consistent with previous findings that AUD and neurodegenerative diseases share some genetic liability (50). Nervous system development related GOBPs were significant (69 genes), and genes involved may predispose to AUD via mechanisms yet to be discovered. GOBPs such as protein metabolic process, cell adhesion, cell development, cell junction organization, movement of cell or subcellular component, cell-cell signaling, and regulation of signaling were also significant. Intuitively, these GOBPs may not seem to be AUD-related, however, among 148 genes only involved in these processes, 20 of them were reported in previous GWAS of AUD related phenotypes with some of them, e.g. *FTO, PDE4B*, and *SLC39A8*, being genome-wide significant in recent large-scale GWAS of AUD (7). In addition, there were seven genes (*EHBP1, EYS, FNBP4, LOC100507053, TNRC6A, WDR7*, and *ZNF462*) that were not involved in any significant GOBPs but were reported by previous GWAS of AUD-related phenotypes. Further studies are needed to elucidate the roles of these genes in predisposing to AUD. By searching the drug target gene database, we found four genes (*DRD2, PDE4B, GRM5*, and *SLC6A9*) are already targets of AUD treatment drugs (**Table S4**). We also found 22 genes are targets of drugs to treat other diseases (**Table S4**) which could be examined and/or repurposed to treat AUD. Studies have found that gene-targeted drugs were more likely to get FDA approval (29, 51, 52), therefore, identifying the roles of genes used in calculating PRS_gene_ could facilitate the development of novel treatment methods.

This study has several limitations. First, while limiting variants to within gene boundaries can help pinpoint AUD genes, many variants with larger effects are intergenic. As a result, the performance of PRS_gene_ was not comparable to those reported previously in EA samples (Lai et al, accepted) and cannot be used to evaluate AUD risks. Second, even variants in a gene may act on other distal genes, therefore, it is possible that these intragenic variants were indexing disease-attributable effects of distal genes. Third, we limited to variants that have effects in both AA and EA, thus, variants that have effects in AA or EA only were excluded, further reducing the performance of PRS_gene_. Fourth, we used posterior effects estimated from the meta-analysis of AA-MVP and EA-PAU. As the EA-PAU had a much larger sample size (>7 times of sample size of AA-MVP), more weight was put on effects estimated from the EA samples. Therefore, for those variants that have different effects between AA and EA, effects from the meta-analysis were biased toward the EA GWAS. Fifth, PRS-CSx used African samples from the 1000 Genomes project as the LD reference panel and the LD structure within that population does not exactly correspond with the AAs in our samples.

In summary, using 858 variants from 410 genes, we calculated gene-based PRS that were significantly associated with AUD in both AA and EA and outperformed PRS calculated using all variants in AA. These 410 genes were enriched in GO biological processes related to ethanol oxidation and synaptic functions, and many of them are likely to cause AUD and are potential drug targets to treat AUD. Our study outlines a novel strategy for PRS calculation in admixed populations and opportunities for new insight into AUD genetics.

## Data Availability

All data produced in the present study are available upon reasonable request to the authors

## ACKNOWLEDGMENTS

**COGA:** The Collaborative Study on the Genetics of Alcoholism (COGA), Principal Investigators B. Porjesz, V. Hesselbrock, T. Foroud; Scientific Director, A. Agrawal; Translational Director, D. Dick, includes eleven different centers: University of Connecticut (V. Hesselbrock); Indiana University (H.J. Edenberg, T. Foroud, Y. Liu, M.H. Plawecki); University of Iowa Carver College of Medicine (S. Kuperman, J. Kramer); SUNY Downstate Health Sciences University (B. Porjesz, J. Meyers, C. Kamarajan, A. Pandey); Washington University in St. Louis (L. Bierut, J. Rice, K. Bucholz, A. Agrawal); University of California at San Diego (M. Schuckit); Rutgers University (J. Tischfield, R. Hart, J. Salvatore); The Children’s Hospital of Philadelphia, University of Pennsylvania (L. Almasy); Virginia Commonwealth University (D. Dick); Icahn School of Medicine at Mount Sinai (A. Goate, P. Slesinger); and Howard University (D. Scott). Other COGA collaborators include: L. Bauer (University of Connecticut); J. Nurnberger Jr., L. Wetherill, X., Xuei, D. Lai, S. O’Connor, (Indiana University); G. Chan (University of Iowa; University of Connecticut); D.B. Chorlian, J. Zhang, P. Barr, S. Kinreich, G. Pandey (SUNY Downstate); N. Mullins (Icahn School of Medicine at Mount Sinai); A. Anokhin, S. Hartz, E. Johnson, V. McCutcheon, S. Saccone (Washington University); J. Moore, Z. Pang, S. Kuo (Rutgers University); A. Merikangas (The Children’s Hospital of Philadelphia and University of Pennsylvania); F. Aliev (Virginia Commonwealth University); H. Chin and A. Parsian are the NIAAA Staff Collaborators. We continue to be inspired by our memories of Henri Begleiter and Theodore Reich, founding PI and Co-PI of COGA, and also owe a debt of gratitude to other past organizers of COGA, including Ting-Kai Li, P. Michael Conneally, Raymond Crowe, and Wendy Reich, for their critical contributions. This national collaborative study is supported by NIH Grant U10AA008401 from the National Institute on Alcohol Abuse and Alcoholism (NIAAA) and the National Institute on Drug Abuse (NIDA).

This study was made possible, in part, with support from the Indiana Clinical and Translational Sciences Institute funded, in part by Award Number UL1TR002529 from the National Institutes of Health, National Center for Advancing Translational Sciences, Clinical and Translational Sciences Award. The content is solely the responsibility of the authors and does not necessarily represent the official views of the National Institutes of Health.

The authors acknowledge the Indiana University Pervasive Technology Institute for providing [HPC (Big Red II, Karst, Carbonate), visualization, database, storage, or consulting] resources that have contributed to the research results reported within this paper.

AA acknowledges K02DA032573, R01DA054869

## CONFILICT OF INTEREST

All authors have no potential conflicts of interest.

## Figure legends

Figure 1. X-axis is each decile and Y-axis is the standardized PRS_gene_.

